# Real-world safety profile of Enfortumab Vedotin: A comprehensive pharmacovigilance analysis based on the FDA Adverse Event Reporting System (FAERS)

**DOI:** 10.64898/2026.06.06.26355060

**Authors:** Qinchuan Xu, Shue Wang, Hong Sun, Xiaoxia Wei, Jiangming Zhong, JiaQin Cai

**Author notes:** **Corresponding author:** (JC). These authors contributed equally to this work (Qinchuan Xu, Shue Wang, JiaQin Cai).

## Abstract

**Background:** This study aimed to evaluate real-world adverse event (AE) signals of EV to provide evidence-based guidance for its safe clinical application.

**Methods:** Data from the FDA Adverse Event Reporting System (FAERS) database from the period of 2019 Q1–2025 Q3 were analyzed. Disproportionality analysis algorithms, including the reporting odds ratio (ROR), proportional reporting ratio (PRR), Bayesian confidence propagation neural network (BCPNN), and empirical Bayes geometric mean (EBGM), were utilized to mine safety signals.The time to onset (TTO) was evaluated using the Weibull distribution model.

**Results:** Among 11,697,906 reports, 4,177 EV-treated patients experienced 14,511 AEs. The most common System Organ Classes (SOCs) were skin and subcutaneous tissue disorders (18.23%), general disorders and administration site conditions (13.17%).Multi-algorithm consensus identified 179 positive signals. Alongside known toxicities (rash, peripheral neuropathy, hyperglycemia), potential new signals emerged, including dysgeusia, atypical skin lesions, and myelosuppression. Median TTO was 14 days, with the Weibull β of 0.736, confirming an “early failure” profile. Subgroup analysis revealed toxicity heterogeneity: patients aged ≥65 and females exhibited stronger signals for fatal severe cutaneous adverse reactions, while patients aged < 65 and males showed higher susceptibility to neurological and metabolic toxicities.

**Conclusions:** The real-world safety profile of EV confirms known toxicities, reveals new risks (e.g., dysgeusia), and shows toxicity concentrated in the first treatment cycle. Clinical practice requires proactive monitoring during the first two weeks using demographic-specific strategies: vigilance for fatal skin toxicity in elderly and female patients, and close follow-up of neurological and metabolic indicators in younger and male populations.

## 1. Introduction

In 2023, bladder cancer (BC) was the seventh most common cancer in the United States, with approximately 82,290 new cases and an estimated 16,710 deaths[1]. By 2024, BC became the sixth most common malignant tumor, accounting for 83,190 new cases and 16,840 deaths [2]. In 2025, it continued to rank as the sixth most common malignant tumor, with around 84,870 new cases and an estimated 17,420 deaths[3]. Clearly, both new cases and deaths from bladder cancer are increasing annually. The majority of BC are non-muscle-invasive bladder cancer (NMIBC), which can be effectively managed in most patients following long-term cystoscopic surveillance, therapeutic interventions, and surgical resection[4]. However, for patients with muscle-invasive bladder cancer (MIBC), despite receiving comprehensive multimodal therapy centered on radical cystectomy—including neoadjuvant chemotherapy or immunotherapy—approximately 45% to 53% still experience disease recurrence. Of recurrence patterns, local recurrences account for only 10% to 30% of cases, whereas the vast majority (70% to 90%) manifest as distant metastases, most commonly involving regional lymph nodes, the lungs, liver, and bones[5]. Morever, BC significantly impairs patients’ quality of life, and imposes a substantial economic burden due to the high cost of treatment [6, 7].

Historically, cisplatin-based regimens have been the standard first-line chemotherapy for locally advanced or metastatic urothelial carcinoma (la/mUC), achieving an objective response rate (ORR) of approximately 49.4% and a median progression-free survival (PFS) of around 7.7 months [8]. BC predominantly affects the elderly population, who frequently present with a physiological decline in renal function. Owing to the significant nephrotoxicity and reliance on renal excretion of cisplatin, approximately 30% to 50% of patients are deemed “cisplatin-unfit” due to renal impairment, and are therefore unable to benefit from standard first-line chemotherapy [9].

In recent years, the emergence of EV, an ADC, has broken this stalemate, receiving its initial global approval from the FDA in December 2019. EV is an ADC targeting Nectin-4; it precisely targets tumor cells via an anti-Nectin-4 monoclonal antibody. Once internalized, the linker is cleaved by proteases within the cell, releasing the cytotoxic payload monomethyl auristatin E (MMAE). This process inhibits microtubule polymerization, subsequently inducing G2/M phase arrest and apoptosis in tumor cells [10]. Results from the Phase III clinical trial (EV-302/KEYNOTE-A39) conducted by Powles T et al. demonstrated that the combination of EV and pembrolizumab achieved a median overall survival (OS) of 31.5 months. This nearly doubles the 16.1 months observed with traditional platinum-based chemotherapy, corresponding to a 53% reduction in the risk of death (HR=0.47, P<0.001)[11]. Building upon these findings, the 2024 NCCN guidelines, as well as the 2025 EAU and CSCO guidelines, have issued major updates, unanimously recommending the EV combination regimen as the preferred first-line standard of care for la/mUC [5, 12, 13].

Previous studies on EV have been conducted under controlled conditions. However, constrained by limited sample sizes and relatively short follow-up periods, these trials may have overlooked various AEs [14]. Therefore, large-scale risk surveillance using the US FAERS, the world’s largest spontaneous reporting system, holds significant clinical value. Based on real-world data from the first quarter of 2019 to the third quarter of 2025, this study aimed to systematically mine and evaluate the safety signals of EV by integrating four statistical methods: ROR, PRR, BCPNN, and EBGM. By accurately profiling the TTO of AEs and conducting multidimensional subgroup analyses across sex, age, and severity, this research seeks to identify potential or unexpected safety risks. Ultimately, these insights will provide a robust scientific basis for clinicians to optimize EV risk management strategies and ensure patient safety.

## 2. Methods

### 2.1 Data collection

Data for this study were obtained from the publicly accessible and authorized FAERS database. The search period spanned from the first quarter of 2019 to the third quarter of 2025, using the drug name “Enfortumab Vedotin” as the search term to capture the most recent data on EV-related adverse reactions. The database is updated quarterly and consists of seven distinct datasets: demographic and administrative information (DEMO), drug information (DRUG), adverse drug reaction information (REAC), patient outcome information (OUTC), information on report sources (RPSR), drug therapy start and end dates (THER), and indications for use/diagnosis (INDI). After removing 1,874,710 duplicate records, a total of 9,823,196 unique reports were retained for subsequent analysis. Data processing and statistical analysis were conducted using SAS 9.4. Data were accessed for research purposes on January 23, 2026, and the authors did not have access to any information that could identify individual participants during or after data collection.

### 2.2 Data extraction and analysis

To ensure data quality, we strictly followed the FDA-recommended deduplication procedure. Specifically, the PRIMARYID, CASEID, and FDA_DT fields were selected from the DEMO table, and the records were sorted by CASEID, FDA_DT, and PRIMARYID in that order. For reports with identical CASEIDs, only the record with the largest FDA_DT field value was retained; if both CASEID and FDA_DT field values were identical, the record with the largest PRIMARYID field value was retained. For drug identification and standardization, the Medical Dictionary for Regulatory Activities (MedDRA) terminology (version 28.1) was used to standardize the drug name as “Enfortumab Vedotin”. To enhance the specificity of signal detection, only AE records where the drug was classified as the “Primary Suspect” (PS) were extracted. All filtered AEs were accurately mapped to the SOC and Preferred Term (PT) levels according to MedDRA terminology for subsequent disproportionality analysis and signal evaluation. The onset time was calculated from the onset date of AE (EVENT_DT field in the DEMO file) minus the start date of enfortumab vedotin treatment (START_DT field in the THER file). Reports with input errors (inaccurate entries, missing data, and EVENT_DT earlier than START_DT) were excluded.

An AE is defined as any untoward medical occurrence in a patient administered a pharmaceutical product, which does not necessarily have a causal relationship with this treatment [15]. To ensure the robustness of signal screening and reduce the risk of false positives, this study simultaneously employed multiple disproportionality analysis methods [16–18], including the ROR, PRR,BCPNN, and EBGM, to detect EV-related adverse event signals. Each method possesses unique advantages: 1) ROR can correct biases caused by a small number of reports for certain events; 2) PRR has higher specificity compared to ROR; 3) BCPNN excels at integrating multi-source data and performing cross-validation; 4) EBGM is capable of detecting signals for rare events. By combining these algorithms, this study aims to leverage their respective advantages to broaden the detection scope, validate the results from multiple perspectives, and rationally utilize their distinct characteristics. The combined application of multiple algorithms allows for cross-validation to reduce false-positive results and detect more potential rare adverse drug reactions. All algorithms rely on a 2×2 contingency table for analysis (S1 Table), and calculations were performed strictly according to the formulas and positive thresholds listed in S2 Table. Higher values obtained through these algorithms indicate stronger signal strength, suggesting a more robust association between EV and the AEs. Furthermore, this study conducted a detailed comparison between the cleaned positive signals and the drug label to identify potential new adverse reaction signals not listed on the label. The initial data cleaning and subsequent statistical modeling for EV were primarily conducted using Microsoft Excel 2024 and R Studio software; the specific technical roadmap is detailed in the flowchart (Fig 1).

**Fig 1.**
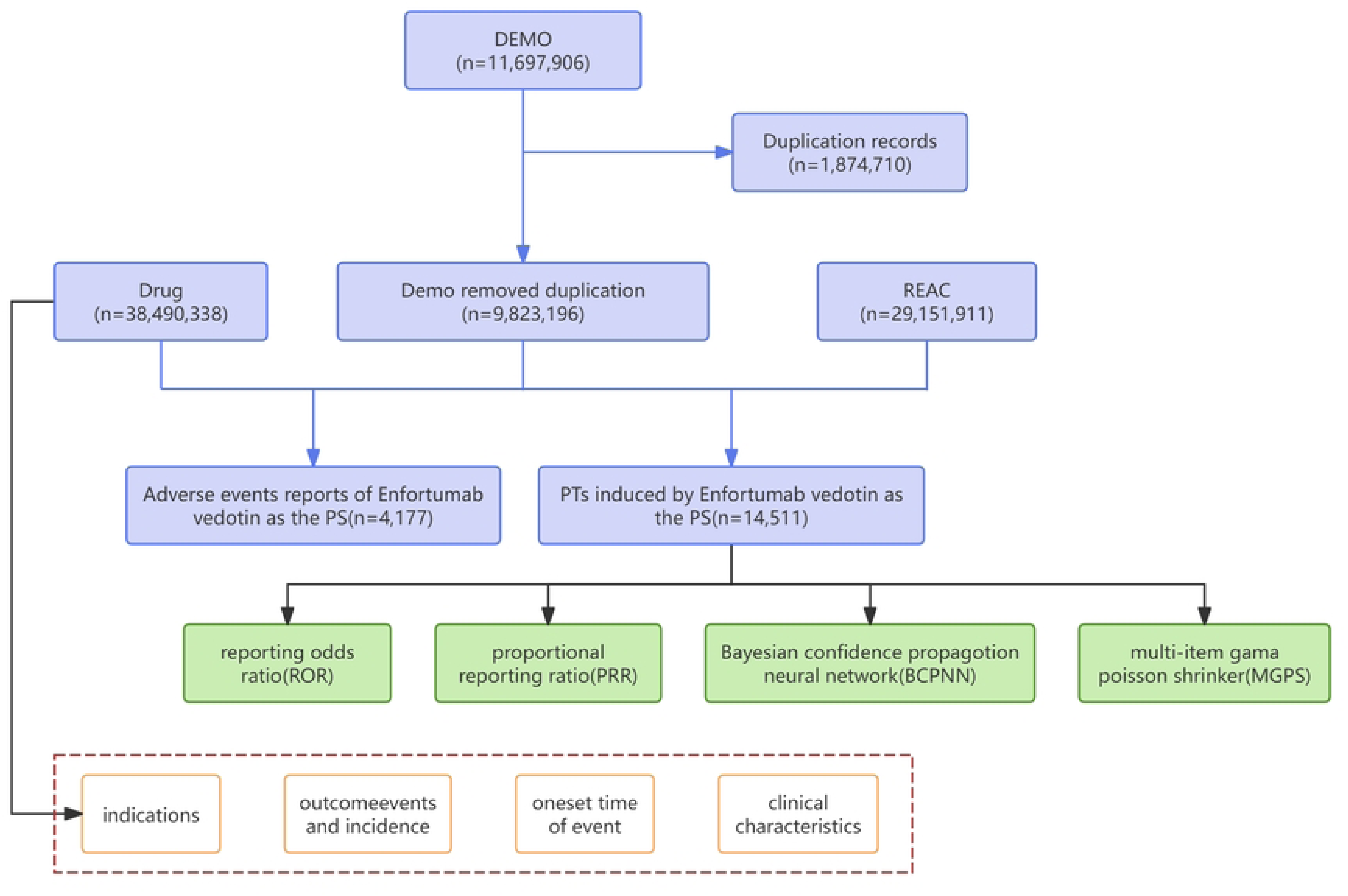
Flowchart of the selection process for EV-related AEs from the FAERS database.

## 3. Results

### 3.1 Basic characteristics of EV-related AEs

From Q1 2019 to Q3 2025, the FAERS database contained 11,697,906 adverse event reports, of which 14,511 reports identified EV as the primary suspect drug, involving 4,177 patients. The number of reports increased annually. Male patients accounted for the majority (3,020, 72.30%), compared to 987 females (23.63%). Most patients were aged ≥65 years (2,484 cases, 59.47%). Physicians were the primary reporters (2,052 cases, 49.13%), followed by consumers (1,080, 25.86%) and pharmacists (993, 23.77%). The main reporting countries were Japan (36.94%) and the United States (34.00%). Regarding clinical outcomes, other serious medical events were most common (2,823 cases, 67.58%), followed by hospitalization (1,444 cases, 34.57%). Detailed information is provided in Table 1.

**Table 1.** Basic information on AEs related to enfortumab vedotin from the FAERS database.

| Variable | Total |
| --- | --- |
| **Number of events** | 4177 |
| **Year** |  |
| 2020 | 231(5.53) |
| 2021 | 316(7.57) |
| 2022 | 457(10.94) |
| 2023 | 848(20.30) |
| 2024 | 982(23.51) |
| 2025 | 1343(32.15) |
| **Sex** |  |
| Female | 987(23.63) |
| Male | 3020(72.30) |
| Unknown | 170(4.07) |
| **Age** |  |
| <65 | 638(15.27) |
| ≥65 | 2484(59.47) |
| Unknown | 1055(25.26) |
| **Reporter** |  |
| Physician | 2052(49.13) |
| Consumer | 1080(25.86) |
| Pharmacist | 993(23.77) |
| Lawyer | 2(0.05) |
| Unknown | 50(1.20) |
| **Reporterd countries** |  |
| Japan | 1543(36.94) |
| United States | 1420(34.00) |
| Canada | 278(6.66) |
| France | 163(3.90) |
| Germany | 73(1.75) |
| Other | 700(16.76) |
| **Outcomes** |  |
| Other serious | 2823(67.58) |
| Hospitalization | 1444(34.57) |
| Death | 799(19.13) |
| Life threatening | 190(4.55) |
| Disability | 50(1.20) |
| Required Intervention to Prevent Permanent Impairment/Damage | 3(0.07) |

Abbreviations: AEs, Adverse events; FAERS, FDA Adverse Event Reporting System.

### 3.2 Signal Detection Results Associated with EV

#### 3.2.1 Signal detection based on the SOC level

Signal detection at the SOC level revealed that ADEs induced by EV encompassed 26 SOCs. Among them, the three most commonly affected systems were skin and subcutaneous tissue disorders (2,646 reports, ROR 3.58, PRR 3.11, IC 1.64, EBGM 3.11), metabolism and nutrition disorders (903 reports, ROR 3.32, PRR 3.18, IC 1.67, EBGM 3.17), and blood and lymphatic system disorders (624 reports, ROR 2.59, PRR 2.52, IC 1.33, EBGM 2.52), indicating strong correlations across all four algorithms. These findings are consistent with the common adverse reactions listed in the drug label, thereby further enhancing the credibility of the data. However, certain SOCs presenting noteworthy adverse reactions, such as psychiatric disorders (139 reports, ROR 0.18, PRR 0.18, IC -2.43, EBGM 0.18) and immune system disorders (46 reports, ROR 0.26, PRR 0.26, IC -1.93, EBGM 0.26), have not yet been documented in the drug label (Table 2).

**Table 2.**
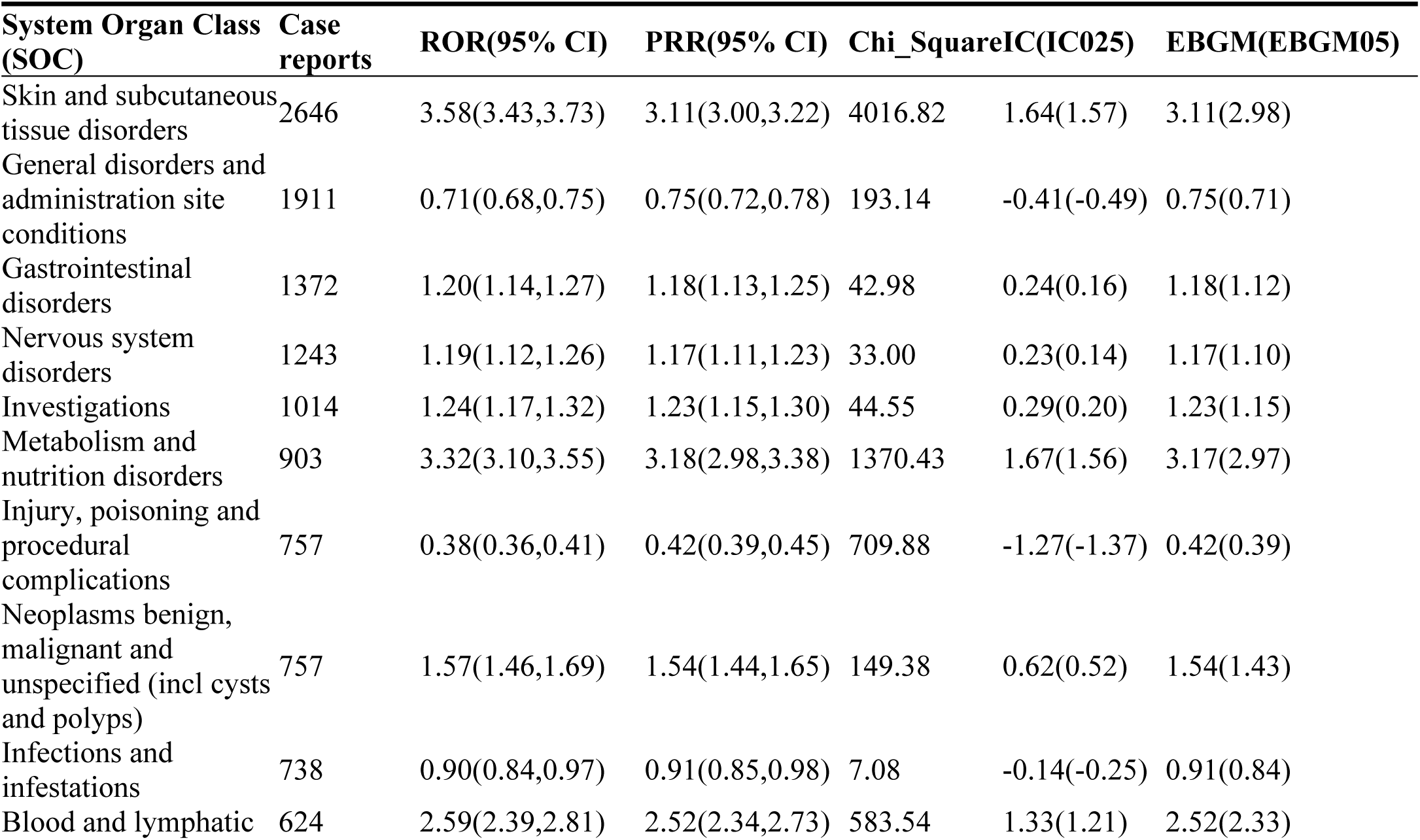
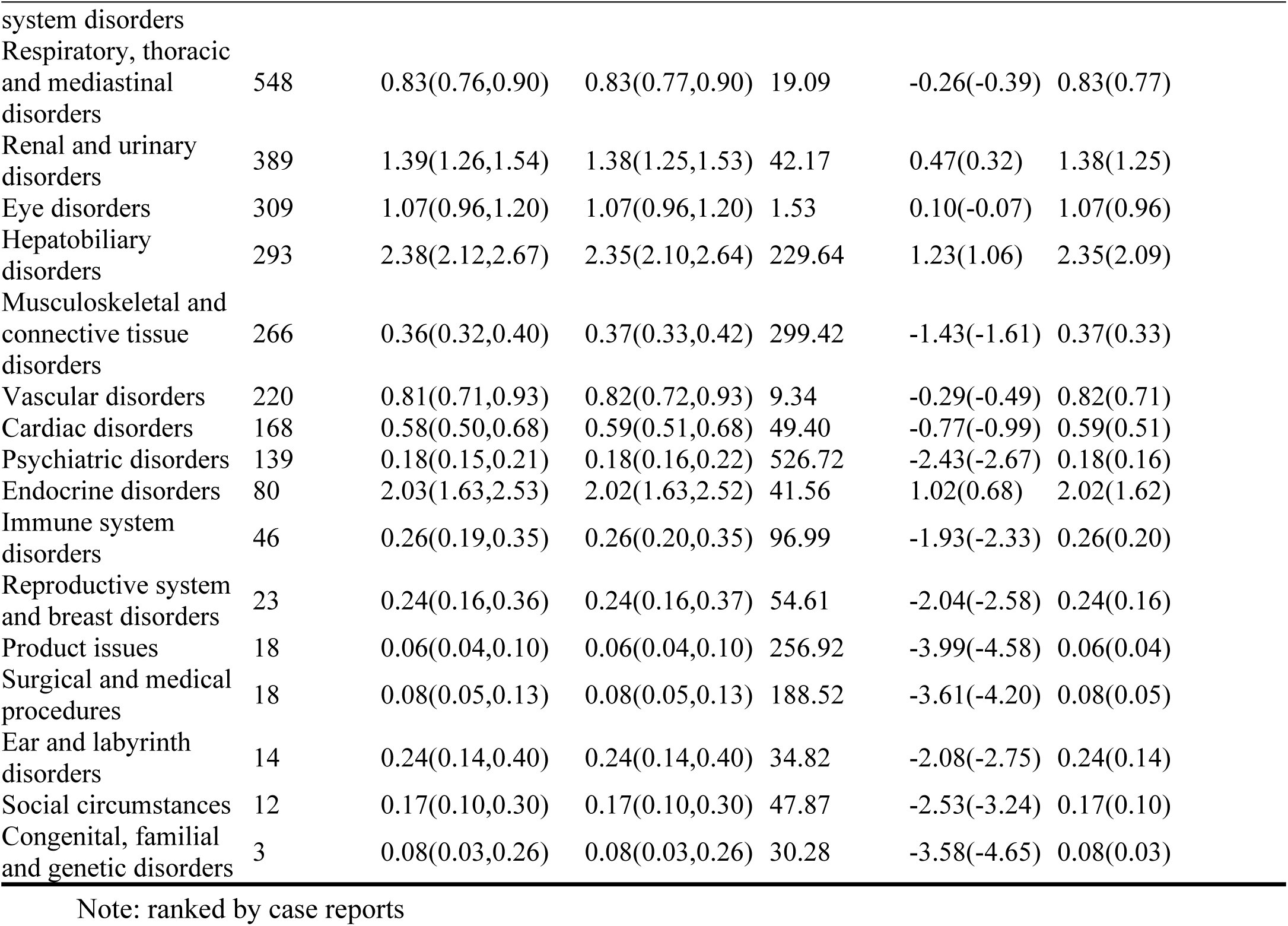
Signal strength of ADEs at the SOC level in FAERS database.

| System Organ Class (SOC) | Case reports | ROR(95% CI) | PRR(95% CI) | Chi_Square | IC(IC025) | EBGM(EBGM05) |
| --- | --- | --- | --- | --- | --- | --- |
| Skin and subcutaneous tissue disorders | 2646 | 3.58(3.43,3.73) | 3.11(3.00,3.22) | 4016.82 | 1.64(1.57) | 3.11(2.98) |
| General disorders and administration site conditions | 1911 | 0.71(0.68,0.75) | 0.75(0.72,0.78) | 193.14 | -0.41(-0.49) | 0.75(0.71) |
| Gastrointestinal disorders | 1372 | 1.20(1.14,1.27) | 1.18(1.13,1.25) | 42.98 | 0.24(0.16) | 1.18(1.12) |
| Nervous system disorders | 1243 | 1.19(1.12,1.26) | 1.17(1.11,1.23) | 33.00 | 0.23(0.14) | 1.17(1.10) |
| Investigations | 1014 | 1.24(1.17,1.32) | 1.23(1.15,1.30) | 44.55 | 0.29(0.20) | 1.23(1.15) |
| Metabolism and nutrition disorders | 903 | 3.32(3.10,3.55) | 3.18(2.98,3.38) | 1370.43 | 1.67(1.56) | 3.17(2.97) |
| Injury, poisoning and procedural complications | 757 | 0.38(0.36,0.41) | 0.42(0.39,0.45) | 709.88 | -1.27(-1.37) | 0.42(0.39) |
| Neoplasms benign, malignant and unspecified (incl cysts and polyps) | 757 | 1.57(1.46,1.69) | 1.54(1.44,1.65) | 149.38 | 0.62(0.52) | 1.54(1.43) |
| Infections and infestations | 738 | 0.90(0.84,0.97) | 0.91(0.85,0.98) | 7.08 | -0.14(-0.25) | 0.91(0.84) |
| Blood and lymphatic | 624 | 2.59(2.39,2.81) | 2.52(2.34,2.73) | 583.54 | 1.33(1.21) | 2.52(2.33) |

| System Organ Class (SOC) | Case reports | ROR(95% CI) | PRR(95% CI) | Chi_Square | IC(0.025) | EBGM(EBGM05) |
| --- | --- | --- | --- | --- | --- | --- |
| system disorders |  |  |  |  |  |  |
| Respiratory, thoracic and mediastinal disorders | 548 | 0.83(0.76,0.90) | 0.83(0.77,0.90) | 19.09 | -0.26(-0.39) | 0.83(0.77) |
| Renal and urinary disorders | 389 | 1.39(1.26,1.54) | 1.38(1.25,1.53) | 42.17 | 0.47(0.32) | 1.38(1.25) |
| Eye disorders | 309 | 1.07(0.96,1.20) | 1.07(0.96,1.20) | 1.53 | 0.10(-0.07) | 1.07(0.96) |
| Hepatobiliary disorders | 293 | 2.38(2.12,2.67) | 2.35(2.10,2.64) | 229.64 | 1.23(1.06) | 2.35(2.09) |
| Musculoskeletal and connective tissue disorders | 266 | 0.36(0.32,0.40) | 0.37(0.33,0.42) | 299.42 | -1.43(-1.61) | 0.37(0.33) |
| Vascular disorders | 220 | 0.81(0.71,0.93) | 0.82(0.72,0.93) | 9.34 | -0.29(-0.49) | 0.82(0.71) |
| Cardiac disorders | 168 | 0.58(0.50,0.68) | 0.59(0.51,0.68) | 49.40 | -0.77(-0.99) | 0.59(0.51) |
| Psychiatric disorders | 139 | 0.18(0.15,0.21) | 0.18(0.16,0.22) | 526.72 | -2.43(-2.67) | 0.18(0.16) |
| Endocrine disorders | 80 | 2.03(1.63,2.53) | 2.02(1.63,2.52) | 41.56 | 1.02(0.68) | 2.02(1.62) |
| Immune system disorders | 46 | 0.26(0.19,0.35) | 0.26(0.20,0.35) | 96.99 | -1.93(-2.33) | 0.26(0.20) |
| Reproductive system and breast disorders | 23 | 0.24(0.16,0.36) | 0.24(0.16,0.37) | 54.61 | -2.04(-2.58) | 0.24(0.16) |
| Product issues | 18 | 0.06(0.04,0.10) | 0.06(0.04,0.10) | 256.92 | -3.99(-4.58) | 0.06(0.04) |
| Surgical and medical procedures | 18 | 0.08(0.05,0.13) | 0.08(0.05,0.13) | 188.52 | -3.61(-4.20) | 0.08(0.05) |
| Ear and labyrinth disorders | 14 | 0.24(0.14,0.40) | 0.24(0.14,0.40) | 34.82 | -2.08(-2.75) | 0.24(0.14) |
| Social circumstances | 12 | 0.17(0.10,0.30) | 0.17(0.10,0.30) | 47.87 | -2.53(-3.24) | 0.17(0.10) |
| Congenital, familial and genetic disorders | 3 | 0.08(0.03,0.26) | 0.08(0.03,0.26) | 30.28 | -3.58(-4.65) | 0.08(0.03) |

Note: ranked by case reports

#### 3.2.2. Signal detection based on the PT level

Using four disproportionality algorithms (ROR, PRR, BCPNN, and EBGM), 179 PT signals meeting the positive thresholds of all four methods were initially identified. After rigorous data cleaning to exclude terms related to EV indications or non-drug-related events [16], the top 30 PTs by case count are presented in Table 3. Notably, peripheral neuropathy (414 reports, ROR 17.84) and hyperglycemia (175 reports, ROR 25.16) showed high reporting frequencies and robust signals. Additionally, several potential new AEs not currently listed on the drug label were identified, including dysgeusia (168 reports, ROR 19.47), skin disorder (165 reports, ROR 20.59), myelosuppression (112 reports, ROR 8.34), pyrexia (190 reports, ROR 2.56), and decreased neutrophil count (71 reports, ROR 6.76). Fig 2 illustrates the disproportionality analysis of the top 30 EV-associated AEs.

**Table 3.** Signal strength of ADEs at the Preferred Terms(PTs) level in FAERS database.

| System Organ Class (SOC) | Preferred Terms (PTs) | Case reports | ROR (95% CI) | PRR (95% CI) | Chi_Square | IC (IC025) | EBGM (EBGM05) |
| --- | --- | --- | --- | --- | --- | --- | --- |
| Skin and subcutaneous tissue disorders | Rash | 598 | 5.86 (5.40,6.36) | 5.66 (5.23,6.12) | 2302.86 | 2.50 (2.36) | 5.64 (5.20) |
| Nervous system disorders | Neuropathy peripheral | 414 | 17.84 (16.17,19.68) | 17.36 (15.78,19.10) | 6338.61 | 4.11 (3.91) | 17.22 (15.61) |
| Skin and subcutaneous tissue disorders | Pruritus | 286 | 3.04 (2.70,3.41) | 3.00 (2.67,3.36) | 382.21 | 1.58 (1.40) | 2.99 (2.66) |
| Metabolism and nutrition disorders | Decreased appetite | 277 | 5.15 (4.58,5.81) | 5.08 (4.52,5.70) | 907.51 | 2.34 (2.14) | 5.06 (4.50) |
| Skin and subcutaneous tissue disorders | Alopecia | 214 | 4.63 (4.05,5.30) | 4.58 (4.01,5.23) | 599 | 2.19 (1.97) | 4.57 (3.99) |
| General disorders and administration site conditions | Pyrexia | 190 | 2.56 (2.22,2.96) | 2.54 (2.21,2.93) | 178.33 | 1.34 (1.12) | 2.54 (2.20) |
| Metabolism and nutrition disorders | Hyperglycaemia | 175 | 25.16 (21.65,29.23) | 24.87 (21.44,28.84) | 3961.62 | 4.62 (4.22) | 24.57 (21.15) |
| Nervous system disorders | Taste disorder | 168 | 19.47 (16.71,22.68) | 19.25 (16.55,22.39) | 2881.59 | 4.25 (3.88) | 19.08 (16.38) |
| Skin and subcutaneous tissue disorders | Skin disorder | 165 | 20.59 (17.65,24.02) | 20.37 (17.49,23.72) | 3009.86 | 4.33 (3.95) | 20.17 (17.29) |
| Blood and lymphatic system disorders | Neutropenia | 120 | 3.29 (2.74,3.93) | 3.27 (2.73,3.90) | 188.88 | 1.71 (1.42) | 3.26 (2.73) |
| Skin and subcutaneous tissue disorders | Stevens-Johnson syndrome | 119 | 35.15 (29.30,42.16) | 34.87 (29.11,41.76) | 3848.74 | 5.10 (4.48) | 34.29 (28.58) |
| Blood and lymphatic system disorders | Myelosuppression | 112 | 8.34 (6.92,10.04) | 8.28 (6.88,9.96) | 714.6 | 3.04 (2.68) | 8.25 (6.85) |
| Blood and lymphatic system disorders | Anaemia | 107 | 2.76 (2.28,3.34) | 2.75 (2.28,3.32) | 119.37 | 1.46 (1.16) | 2.75 (2.27) |
| Nervous system disorders | Hypoaesthesia | 96 | 3.18 (2.60,3.89) | 3.16 (2.59,3.86) | 142.21 | 1.66 (1.33) | 3.16 (2.59) |
| Skin and subcutaneous tissue disorders | Toxic epidermal necrolysis | 88 | 28.52 (23.09,35.22) | 28.35 (22.99,34.97) | 2290.38 | 4.81 (4.11) | 27.97 (22.65) |
| Respiratory, thoracic and mediastinal disorders | Interstitial lung disease | 82 | 7.16 (5.76,8.90) | 7.13 (5.74,8.84) | 430.58 | 2.83 (2.41) | 7.10 (5.72) |
| Blood and lymphatic system disorders | Febrile neutropenia | 82 | 4.97 (4.00,6.18) | 4.95 (3.99,6.14) | 257.92 | 2.30 (1.92) | 4.94 (3.97) |
| Investigations | Neutrophil count decreased | 71 | 6.81 (5.39,8.60) | 6.78 (5.37,8.56) | 349 | 2.76 (2.30) | 6.76 (5.35) |
| Skin and subcutaneous tissue disorders | Skin exfoliation | 71 | 2.94 (2.33,3.72) | 2.93 (2.33,3.70) | 90.45 | 1.55 (1.17) | 2.93 (2.32) |
| Eye disorders | Dry eye | 70 | 5.24 (4.14,6.63) | 5.22 (4.13,6.60) | 238.39 | 2.38 (1.95) | 5.21 (4.12) |
| Metabolism and nutrition disorders | Dehydration | 70 | 2.78 (2.20,3.52) | 2.77 (2.19,3.50) | 79.37 | 1.47 (1.09) | 2.77 (2.19) |
| Hepatobiliary disorders | Hepatic function abnormal | 70 | 8.37 (6.61,10.59) | 8.33 (6.59,10.53) | 450.12 | 3.05 (2.57) | 8.30 (6.56) |
| Gastrointestinal disorders | Dry mouth | 67 | 4.12 (3.24,5.24) | 4.11 (3.24,5.22) | 157.49 | 2.04 (1.62) | 4.10 (3.23) |
| Skin and subcutaneous tissue disorders | Skin toxicity | 65 | 45.03 (35.20,57.61) | 44.84 (35.08,57.30) | 2725.21 | 5.46 (4.37) | 43.88 (34.30) |
| Skin and subcutaneous tissue disorders | Blister | 62 | 4.94 (3.85,6.35) | 4.93 (3.84,6.32) | 193.79 | 2.30 (1.85) | 4.92 (3.83) |
| Skin and subcutaneous tissue disorders | Rash pruritic | 55 | 4.66 (3.57,6.07) | 4.64 (3.57,6.05) | 157.08 | 2.21 (1.74) | 4.64 (3.56) |
| General disorders and administration site conditions | Oedema peripheral | 52 | 2.89 (2.20,3.79) | 2.88 (2.20,3.78) | 63.86 | 1.53 (1.08) | 2.88 (2.19) |
| Skin and subcutaneous tissue disorders | Dermatitis bullous | 52 | 35.51 (26.98,46.73) | 35.38 (26.91,46.52) | 1707.45 | 5.12 (4.01) | 34.79 (26.43) |
| Skin and subcutaneous tissue disorders | Rash erythematous | 51 | 4.73 (3.59,6.22) | 4.71 (3.58,6.20) | 148.96 | 2.23 (1.73) | 4.70 (3.57) |
| Skin and subcutaneous tissue disorders | Skin reaction | 49 | 15.72 (11.86,20.83) | 15.67 (11.84,20.75) | 667.99 | 3.96 (3.18) | 15.56 (11.74) |

Note: ranked by case reports

**Fig 2.**
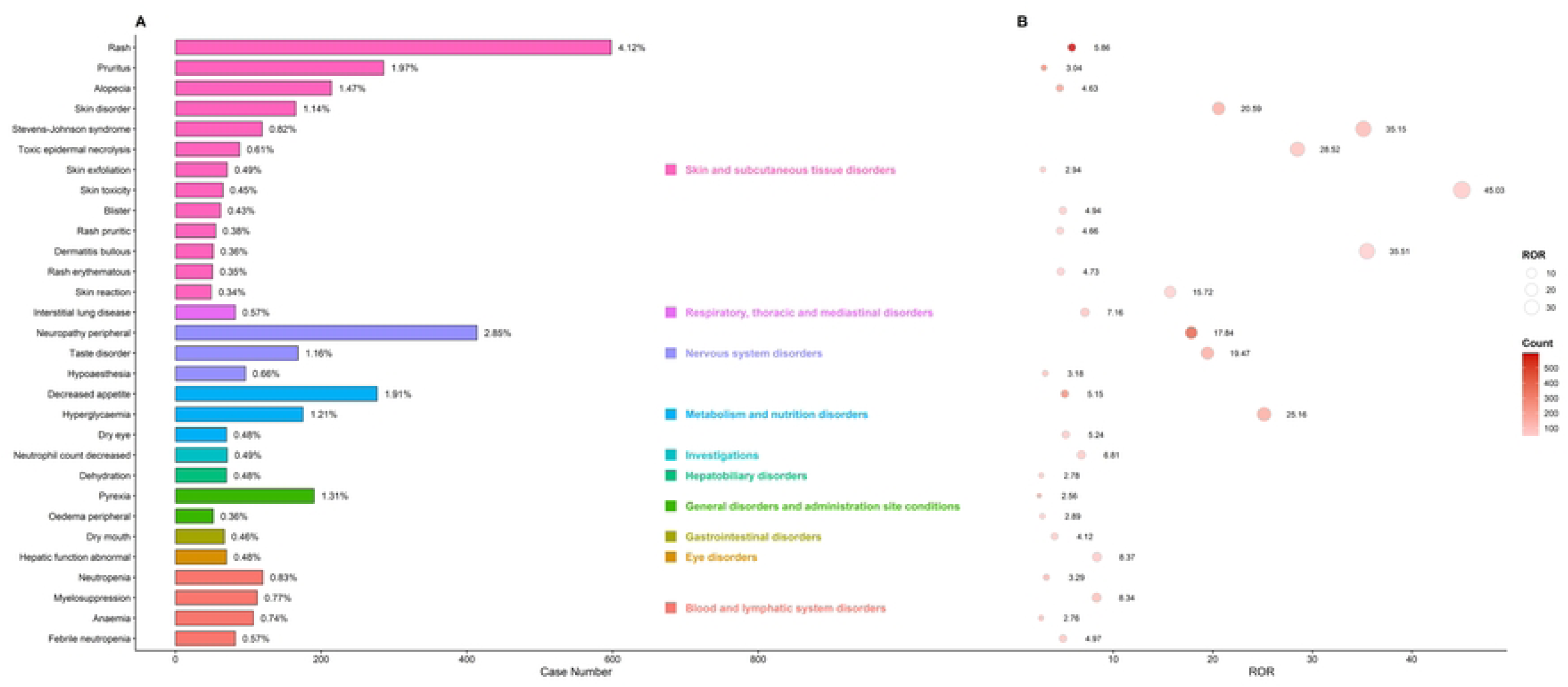
Disproportionality analysis of the top 30 adverse events by case number for EV. (A) The horizontal bar chart displays the report numbers and proportions of the top 30 PTs. The colors and square markers indicate the specific SOC to which each PT belongs. (B) The bubble chart illustrates the corresponding signal strengths based on the ROR. Bubble size is proportional to the magnitude of the ROR, while the color gradient (from light to dark red) reflects the case count.

#### 3.2.3. TTO Analysis of EV-Related AEs

After excluding excluding reports with missing or anomalous onset times, 1,738 cases with valid TTO data were analyzed, revealing a median TTO of 14.00 days (IQR: 7.00–45.00 days). As shown in Fig 3, more than half of the adverse reactions occurred within the first two weeks of EV treatment, indicating an early concentration of events. A Weibull distribution model [17]. fitted to the TTO data yielded a scale parameter (α) of 39.64 (95% CI: 36.91–42.57) and a shape parameter (β) of 0.736 (95% CI: 0.711–0.762). The upper limit of the 95% CI for β was significantly below 1, indicating that the risk of EV-related adverse reactions gradually decreased over time—a typical “early failure” profile. Consistent with this pattern, the majority of adverse events occurred within the first month of treatment[18], with both case numbers and proportions declining steadily thereafter.

**Fig 3.**
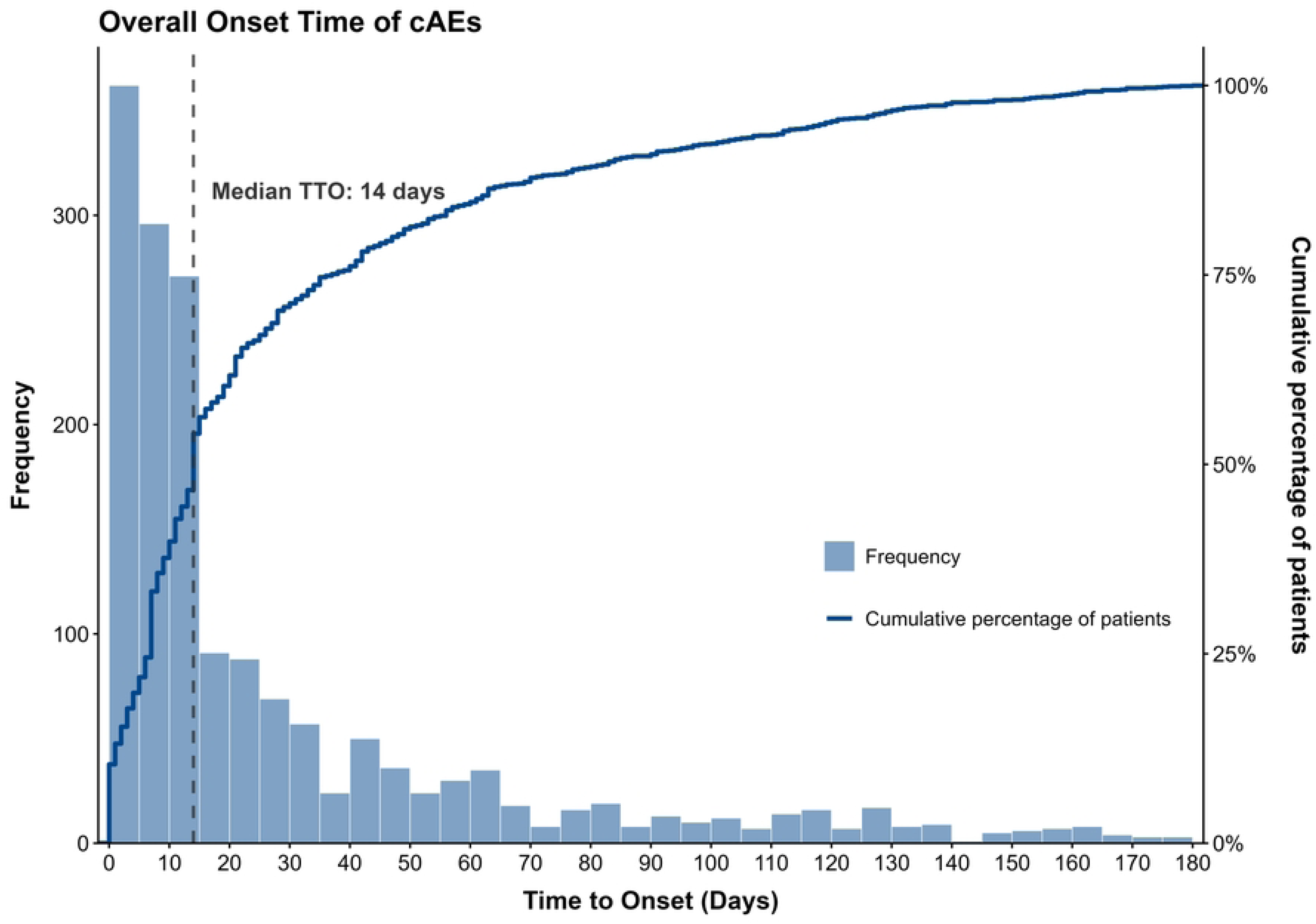
Frequency distribution and cumulative percentage curve of overall time to onset for adverse events. The histogram (left Y-axis) displays the absolute frequency distribution of the onset time (in days). The step curve (right Y-axis) represents the cumulative percentage of adverse events over time. The black dashed line indicates the median time to onset (Median TTO = 14 days).

### 3.3. Subgroup Analysis

#### 3.3.1 Subgroup Analysis by Gender

As shown in Fig 4A, no significant baseline sex difference was observed in overall EV-related adverse reactions. However, male patients accounted for the majority of reports (3,020, 72.30%), compared to 987 (23.63%) for females. This difference reflects the higher incidence of urothelial carcinoma in men.[19].To further explore sex-specific differences, a gender-stratified heatmap based on ROR values was generated(Fig 4B). At the SOC level, notable differences emerged. Female patients exhibited stronger signals for skin and subcutaneous tissue disorders, suggesting greater susceptibility to cutaneous toxicity. In contrast, male patients showed stronger signals for metabolism and nutrition disorders.

**Fig 4.**
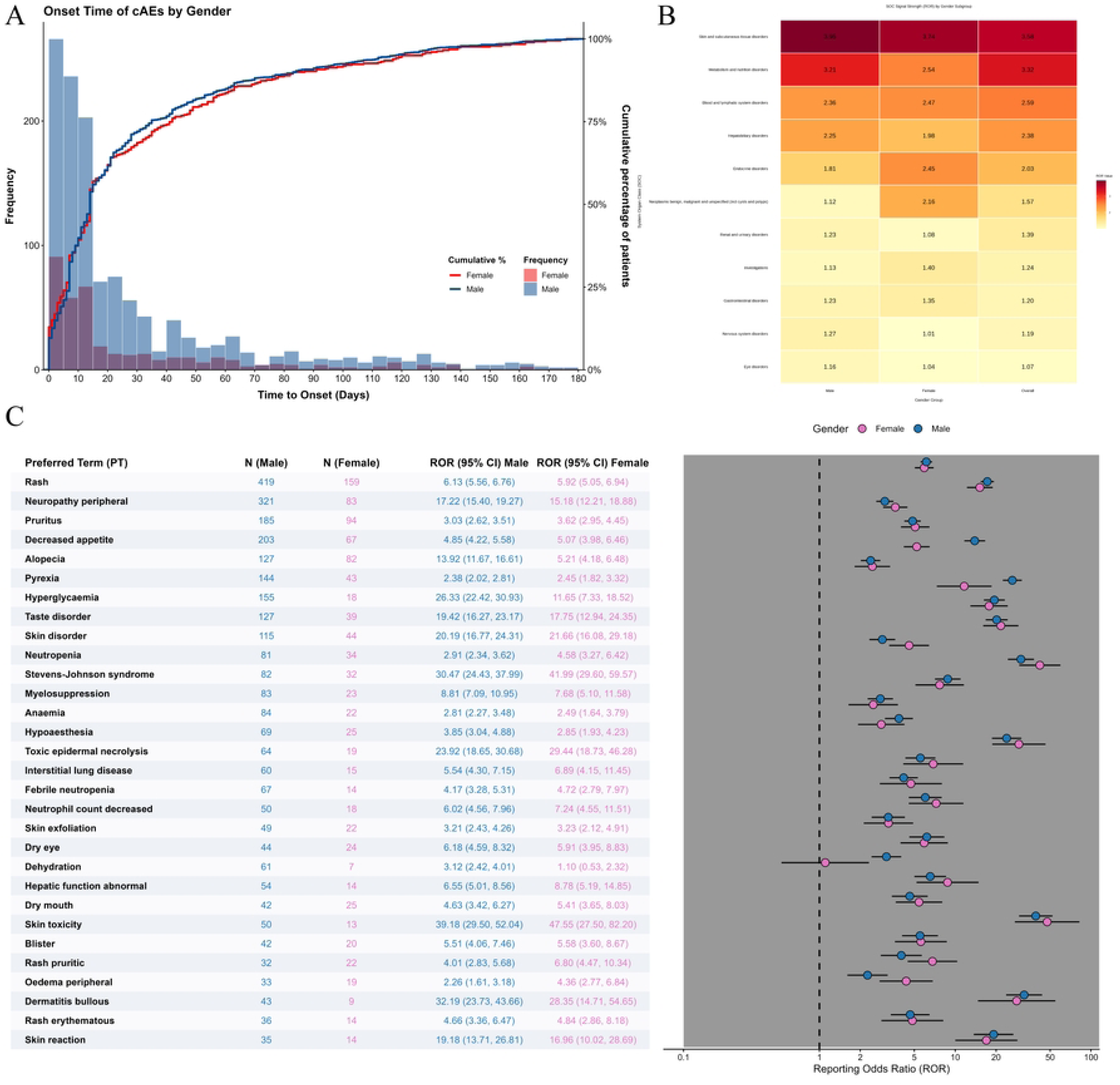
(A)Frequency distribution and cumulative percentage curve of time to onset, stratified by sex. The figure compares the onset time patterns between male (dark blue) and female (red) patients. The overlapping histograms (left Y-axis) show the absolute frequency of adverse events within different time intervals for each gender. The step curves (right Y-axis) illustrate the cumulative percentage of adverse events within the respective gender groups. (B) **Heatmap of ROR for EV-related adverse events at the SOC level, stratified by sex.** The color gradient represents the magnitude of the ROR values: transitions from light yellow to dark red indicate an increasing ROR, corresponding to a stronger association (higher risk) between the drug and the adverse event. The numbers within the heatmap cells represent the specific calculated ROR values. The x-axis displays the gender subgroups (Male; Female; Overall: the entire study population). To highlight the core safety profile, only SOCs demonstrating a positive signal (ROR > 1) in the overall population are displayed. Abbreviations: ROR, Reporting Odds Ratio; SOC, System Organ Class. (C) **Forest plot comparing ROR of the top 30 most frequently reported adverse events, stratified by sex.** This figure illustrates the signal strengths of the top 30 most frequently reported PTs associated with EV across different gender subgroups. Blue dots represent the Male subgroup, and pink dots represent the Female subgroup; error bars indicate the 95% confidence intervals (95% CI). The X-axis is plotted on a logarithmic scale. The vertical dashed line represents the line of no effect (ROR = 1). A signal is considered statistically significant if the lower limit of the 95% CI is > 1 (i.e., the error bar does not cross the dashed line).Abbreviations: PT, Preferred Term; ROR, Reporting Odds Ratio; CI, Confidence Interval; N, Number of reports.

Among the 151 valid PTs (after excluding indication- and non-drug-related events), the top 30 most frequently reported PTs were selected for sex-stratified ROR comparison (Fig 4C). Clear sex disparities were observed. Males showed stronger signals for metabolic disorders and certain skin appendage damages, including hyperglycemia (ROR: 26.33 vs. 11.65 in females) and alopecia (13.92 vs. 5.21). Signals for bullous dermatitis were also more concentrated in males. Conversely, females faced higher risks of severe, life-threatening skin reactions. ROR values for Stevens-Johnson syndrome and toxic epidermal necrolysis were substantially higher in females (41.99 and 29.44, respectively) than in males (30.47 and 23.92). Moreover, the overall skin toxicity signal was markedly stronger in females (ROR 47.55) than in males (ROR 39.18).

#### 3.3.2 Subgroup Analysis by Age

As shown in Fig 5A, EV-related adverse events predominantly occurred in older patients, with the elderly group (≥65 years) accounting for the majority of reports (2,484 cases, 59.47%) compared to the younger group (<65 years, 638 cases, 15.28%), consistent with the higher incidence of urothelial carcinoma in the elderly population[19]. An age-stratified heatmap (Fig 5B) revealed that elderly patients exhibited stronger signals for skin and subcutaneous tissue disorders, while younger patients showed stronger signals for metabolism and nutrition disorders. Further analysis of the top 30 PTs (Fig 5C) demonstrated significant age-related heterogeneity: elderly patients faced higher risks of severe cutaneous adverse reactions, including skin disorder (ROR: 25.00 vs. 13.06), skin toxicity (32.91 vs. 24.84), toxic epidermal necrolysis (TEN,21.21 vs. 12.81), and Stevens-Johnson syndrome (SJS,25.36 vs. 19.25); conversely, younger patients showed stronger signals for neurological, mucosal, and sensory toxicities, such as peripheral neuropathy (16.61 vs. 11.27), dysgeusia (20.07 vs. 11.27), dry eye (8.16 vs. 11.27), and dry mouth (6.72 vs. 11.27).

**Fig 5.**
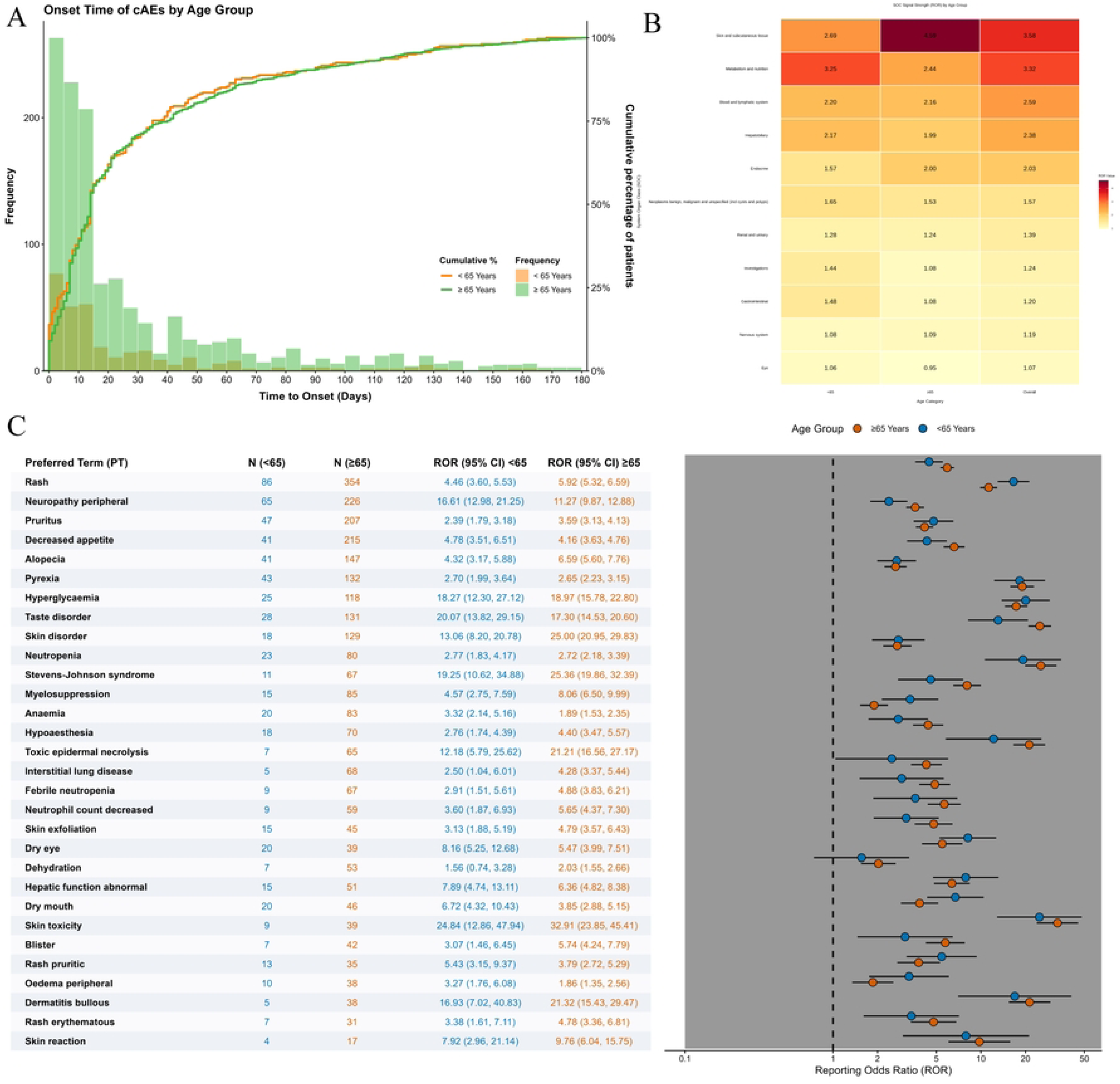
**(A)Frequency distribution and cumulative percentage curve of time to onset, stratified by age.** The figure compares the onset time patterns between the elderly group (≥ 65 years, forest green) and the younger group (< 65 years, bright orange). The overlapping histograms (left Y-axis) display the absolute frequency of adverse events within different time intervals for each age group. The step curves (right Y-axis) illustrate the cumulative percentage of adverse events within the respective age cohorts. (B) **Heatmap of ROR for EV-related adverse events at the SOC level, stratified by age.** The color gradient illustrates the magnitude of the ROR values: a shift from light yellow to dark red denotes an increasing ROR, reflecting progressively stronger signal intensities and higher risks for the respective adverse events. The numeric values inside the cells indicate the exact ROR calculated for each subgroup. The x-axis categorizes patients into different age strata (<65: patients younger than 65 years; ≥65: patients aged 65 years and older; Overall: the entire study population). SOCs without significant risk signals in the overall population were excluded from this visualization. Abbreviations: ROR, Reporting Odds Ratio; SOC, System Organ Class. (C) **Forest plot comparing ROR of the top 30 most frequently reported adverse events, stratified by age.** This figure compares the signal strengths of the top 30 most frequently reported PTs associated with EV across different age subgroups. Blue dots represent the <65 years age group, and orange-red dots represent the ≥65 years age group; error bars indicate the 95% confidence intervals (95% CI). The X-axis is plotted on a logarithmic scale (Log10). The vertical dashed line serves as the reference line (ROR = 1). A risk signal is considered statistically significant if the lower limit of the 95% CI is > 1.Abbreviations: PT, Preferred Term; ROR, Reporting Odds Ratio; CI, Confidence Interval; N, Number of reports.

#### 3.3.3 Subgroup Analysis by Severity

As shown in Fig 6, the majority of EV-related AEs were classified as serious (3,570 cases, 85.47%), while non-serious reports accounted for only 607 cases (14.53%), highlighting that EV treatment for advanced urothelial carcinoma frequently leads to hospitalization or life-threatening conditions. A forest plot comparing RORs between serious and non-serious subgroups for the top 30 PTs (Figure 6, with logarithmic scale) revealed two clinically relevant findings. First, serious reports showed substantially stronger signals for disability-related toxicities, including peripheral neuropathy (ROR: 14.55 vs. 1.73), skin lesion (26.12 vs. 5.26), and dysgeusia (26.91 vs. 15.47), indicating a stronger association with clinically severe cases. Second, extremely high ROR values for severe cutaneous adverse reactions (SCARs), such as TEN (2,766.72) and SJS (351.05), were observed in the non-serious subgroup. This is a known mathematical artifact of spontaneous reporting systems: because SCARs are inherently life-threatening, their background frequency in the non-serious category is nearly zero, and even a few miscoded reports can produce spuriously elevated RORs. In reality, the absolute number of SCAR cases remains concentrated in the serious subgroup.

**Fig 6.**
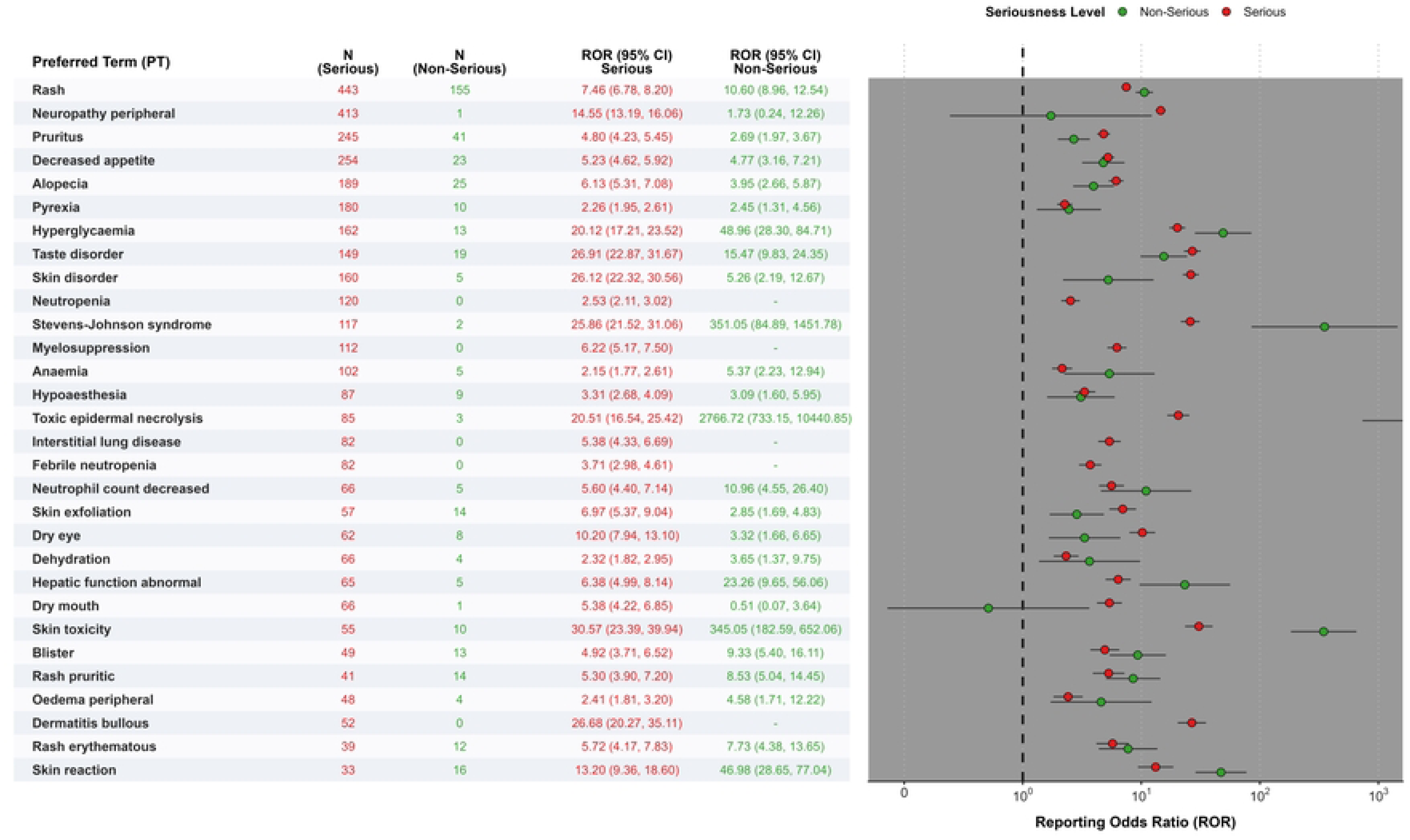
Forest plot comparing ROR of the top 30 most frequently reported adverse events, stratified by severity. This figure displays the differences in signal strengths of the top 30 most frequently reported PTs associated with EV between Serious and Non-Serious report subgroups. Red dots represent Serious reports, and green dots represent Non-Serious reports; error bars indicate the 95% confidence intervals (95% CI). Entries with N=0 or a dash (“-”) for the ROR CI indicate that no reports were recorded for that specific PT in the given subgroup. The X-axis uses a logarithmic scale, and the vertical dashed line is the reference line of no effect (ROR = 1).Abbreviations: PT, Preferred Term; ROR, Reporting Odds Ratio; CI, Confidence Interval; N, Number of reports.

## 4. Discussion

This study represents a comprehensive and in-depth evaluation of the large-sample, real-world safety profile of EV post-marketing. Based on 4,177 target AE reports from the FAERS database between the first quarter of 2019 and the third quarter of 2025, this study comprehensively utilized four data mining algorithms—ROR, PRR, BCPNN, and EBGM—to minimize false-positive rates. More importantly, during the data cleaning stage, this study strictly excluded confounding terms related to EV indications (such as urothelial carcinoma and its progression), ultimately screening out 179 PTs that demonstrated positive signals across all four algorithms. This methodological design not only effectively eliminated the interference of disease progression on drug safety but also ensured that the finally identified safety signals possess extremely high statistical power and clinical reference value. The data indicate that skin and subcutaneous tissue disorders (2,646 reports), as well as general disorders and various nervous system disorders (909 reports), are the primary toxic manifestations of EV.

From the perspective of pharmacological mechanisms, EV is composed of a Nectin-4-directed monoclonal antibody conjugated to the microtubule inhibitor MMAE. Since Nectin-4 is not only highly expressed in urothelial carcinoma but also widely expressed in normal human skin tissues (such as epidermal keratinocytes and hair follicles), this “on-target off-tumor toxicity” is the core mechanism leading to the frequent occurrence of skin and subcutaneous tissue adverse reactions, including rash and pruritus [20, 21]. Furthermore, as the conjugated payload, MMAE exerts a strong microtubule inhibitory effect; its shedding and accumulation around nerve cells can interfere with axonal transport, which biologically aligns with the strong signals for “peripheral neuropathy” observed in this study [22].

A major highlight of this study is that, after excluding known adverse reactions, we utilized the quadruple-algorithm approach to identify several potential new risk signals that have not been explicitly included or adequately warned about in the drug label, particularly dysgeusia and the yet-to-be-fully-defined skin lesion. The frequent occurrence of dysgeusia may have been underestimated in traditional clinical trials. On the one hand, its mechanism may be related to MMAE’s microtubule inhibitory effect involving the cranial nerves responsible for taste conduction (such as the facial and glossopharyngeal nerves), presenting as a specific manifestation of generalized neurotoxicity [22]; on the other hand, it might also be due to the direct cytotoxicity of free MMAE on highly proliferative taste bud cells within the oral mucosa [23]. Additionally, previous literature and FDA safety warnings have clearly stated that EV may induce fatal SJS or TEN [24] [25]. In their early onset stages, these fatal bullous dermatoses often manifest merely as non-specific, atypical skin lesions that are difficult to definitively categorize. Therefore, the large volume of generalized “skin lesion” reports serves as a strong warning to clinicians: any newly developed atypical skin lesions in the early stages of medication (especially within the median onset time of 14 days indicated in this study) should be regarded as prodromal symptoms of potential severe cutaneous reactions, necessitating the most rigorous proactive monitoring and early intervention.

Regarding the TTO characteristics of adverse reactions, the Weibull distribution shape parameter (β value) in this study showed that the overall β value for adverse reactions was 0.73 (95% CI: 0.71–0.76, β < 1), and the median TTO was only 14 days, clearly indicating that EV-related adverse reactions belong to the “early failure type”. In particular, for skin and subcutaneous tissue disorders, which had the highest occurrence frequency, the median TTO was even shorter (only 10 days, β = 0.81). This strongly suggests to clinicians that the majority of severe toxic reactions to EV tend to erupt within the first two treatment cycles (especially the first two weeks) following the initial administration, making this time window a critical period for clinical monitoring.

In terms of demographic distribution, this study found that EV adverse event reports exhibited significant gender and age clustering. Among patients with explicitly recorded baseline information, males comprised 72.30% of the cohort, and elderly patients (≥65 years) reached 59.47% (median age 73 years). This distribution pattern aligns with the epidemiological characteristic of urothelial carcinoma being highly prevalent in elderly males. Elderly patients frequently present with an underlying decline in hepatic and renal functions, slowed metabolism, and multiple concomitant medications, which may lead to a decreased clearance rate of MMAE in vivo, thereby increasing the risk of peripheral neuropathy or severe skin toxicities. Therefore, for the elderly male population aged ≥65 years, a comprehensive baseline assessment of neurological function and skin condition is recommended before initiating EV treatment; during the first 14 days post-medication, close follow-up and prophylactic skin care should be implemented. If early toxicity signals appear, including atypical skin lesions or dysgeusia, intervention strategies such as dose de-escalation or temporary discontinuation should be promptly adopted to prevent the occurrence of severe or irreversible AEs.

This study has several limitations. First, as a spontaneous reporting system, FAERS data are subject to underreporting, misreporting, and bias toward severe or rare events. The “Weber effect” (a surge in reporting during the initial post-marketing period of a new drug) may also confound the analysis. Second, the lack of precise exposure data (i.e., person-years) prevents calculation of true incidence rates; thus, disproportionality analysis can only suggest statistical associations, not establish causality. Third, despite excluding indication- and non-drug-related terms, the absence of detailed patient information, exact dosages, and concomitant medications (e.g., frequent co-administration with immune checkpoint inhibitors) may introduce residual confounding.

Notwithstanding these limitations, this study offers valuable insights. Traditional pre-marketing trials (e.g., EV-301) are often limited by small sample sizes, strict eligibility criteria, and short follow-up durations, failing to capture the complexity of real-world clinical practice. By leveraging the FAERS database and employing four complementary data mining algorithms for cross-validation, this study mitigates the shortcomings of any single method and enhances the robustness of signal detection. These findings provide clinicians with important early warnings for individualized treatment strategies and lay the groundwork for future prospective cohort studies.

This real-world analysis of EV using FAERS data confirms known toxicities and identifies new signals (dysgeusia, atypical skin lesions, myelosuppression). EV toxicity peaks within the first treatment cycle (median 14 days), fitting an “early failure” pattern. Elderly patients and females are more prone to severe skin reactions, while younger patients and males face higher risks of neurological and metabolic toxicities. These findings support early, individualized monitoring strategies to improve patient safety.

## Data Availability

No primary data were generated by this study. The following existing data sources were used: The FDA Adverse Event Reporting System (FAERS) database, which is publicly available from the U.S. Food and Drug Administration (FDA) at https://fis.fda.gov/extensions/FPD-QDE-FAERS/FPD-QDE-FAERS.html.

## Declarations

### Human Ethics declaration

Not applicable. This study used publicly available, anonymized data from the FDA Adverse Event Reporting System (FAERS), which does not constitute human subjects research requiring ethical approval.

### Consent to Participate

Not applicable. Since the study relied on an anonymized, publicly available database (FAERS), informed consent from human participants was not required.

### Consent for publication

Not applicable.

### Data availability

The datasets analyzed during the current study are available in the FDA Adverse Event Reporting System (FAERS) database, which is a publicly accessible repository.

## Acknowledgements

Not applicable.

## Supporting information

**S1 Table.**
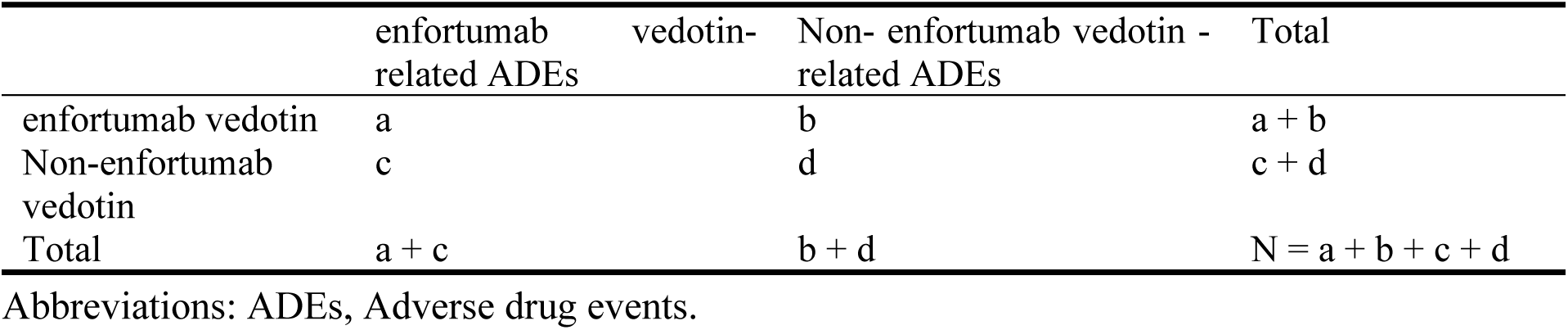
Four grid table.

**S2 Table.**
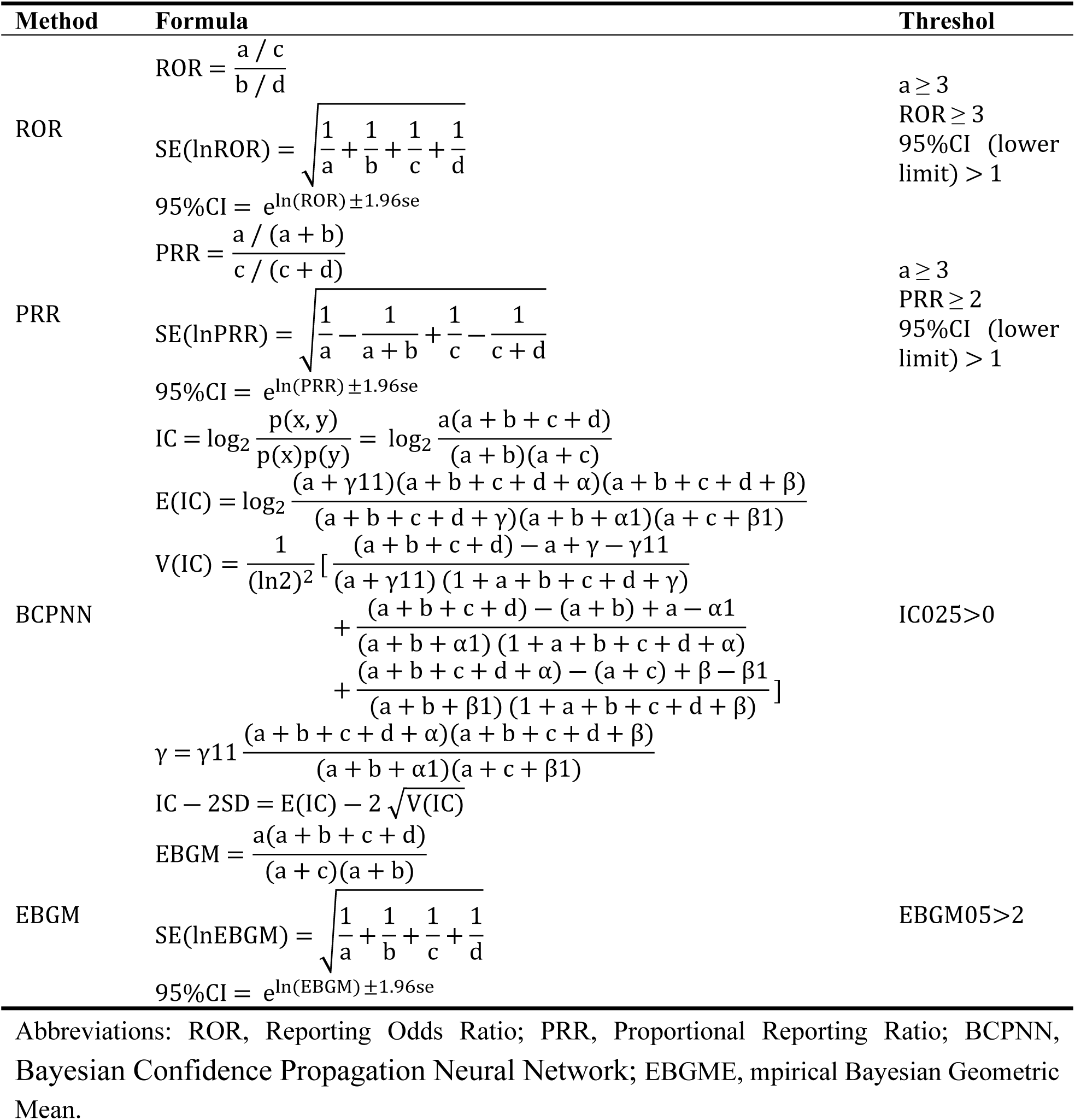
ROR, PRR, BCPNN, and EBGM methods, formulas, and thresholds.

